# Interim Analysis of Dose-Escalated Preoperative Breast Irradiation: A Phase Ib study

**DOI:** 10.1101/2025.08.29.25334741

**Authors:** Ravi Patel, Emilia Diego, Hayeon Kim, Wendie A. Berg, Rohit Bhargava, Hong Wang, Quratulain Sabih, Priscilla McAuliffe, Adam Brufsky, Julia Foldi, Shannon Puhalla, Kevin Kane, Marija Balic, Bob Edinger, Shandong Wu, Heath Skinner, Parul Barry

**Affiliations:** University of Pittsburgh School of Medicine; University of Pittsburgh; University of Pittsburgh Medical Center; UPMC Hillman Cancer Center

## Abstract

**Introduction:** Breast irradiation (RT) is often recommended as part of breast conservative therapy (BCT) and is often delivered daily over multiple weeks after breast conserving surgery (BCS). Information regarding tumor response is lost when BCS is performed before oncologic treatment. Short course partial breast irradiation delivered preoperatively (PRT) is a potential solution.

**Methods:** Following informed consent, ten participants enrolled in one of four cohorts of HCC 22-003 (NCT05464667). Each of the first three cohorts enrolled three participants, and one enrolled in the fourth cohort, with dose-escalation to the gross tumor volume (GTV) by 5 Gy (from 30 to 45 Gy). Participants underwent BCS within five to ten days following PRT. Pre- and post-RT contrast-enhanced breast MRI and blood work was obtained for those who consented and were able to undergo such procedures. Magee Equation 3 was calculated on all core biopsy specimens and genomic oncotype testing performed as indicated per medical oncology expert opinion.

**Results:** No patients experienced ≥ grade 2 or higher acute toxicity. No pathologic complete response (pCR) occurred following PRT on interim analysis of cohorts one to three. Three patients (two of which were in cohort three, 40 Gy to GTV) had decrease in tumor size noted from pre-to post-PRT MRI. Further, two of the three patients in cohort three had substantial pathologic tumoral response to PRT. Significant increase in TNFa and INFb serum levels were seen post-RT for cohort three compared to cohorts one and two.

**Conclusions:** Dose-escalated PRT is safe, feasible, and well tolerated. There appears to be a tumoral response noted on MRI and final pathology with dose escalation to 40 Gy to the GTV. Continuation of dose-escalation to cohort four is planned with final analysis reported following completion.

## Introduction

NSABP B-06 established breast conservative therapy (BCT) as the standard treatment for early-stage breast cancer ^1^. Currently multiple post-operative treatment regimens exist, ranging from 4 weeks of daily whole breast irradiation (RT) to as little as five treatments of partial breast irradiation (PBI) ^2^. Often, PBI for early-stage luminal breast cancers is preferred due to improved toxicity profile and ease of treatment ^3 4^. Additionally, accelerated regimens reduce the financial burden and opportunity costs faced by those receiving longer courses of whole breast irradiation, and lack of compliance and completion in longer treatment courses result in reduced survival ^5,6^. Unfortunately, PBI is only offered in a minority of patients, in part due to challenges with planning in the setting of large post-operative treatment volumes^7^.

Preoperative treatment, delivered in preparation for definitive breast surgery, has primarily been systemic chemotherapy (CT) and endocrine therapy (ET). Such therapy has improved breast conservation rates and has also provided a window into oncologic outcomes based on pathologic response^8^. The role of preoperative systemic chemotherapy in hormone-receptor positive tumors is reduced following the publication of TAILORx^9^, but the need to understand tumor response persists.

Simultaneously, the role of RT relative to CT and/or ET has been reduced: approximately 75% of breast RT trials are evaluating ways to omit RT, in part due to radiation and financial toxicity concerns ^10^. Trials such as CALGB 943^11^, PRIME II^12^, IDEA ^13^and LUMINA ^14^have evaluated omission of RT in favor of ET. Ongoing or recently completed studies such as NRG BR007 (DEBRA, NCT04852887), NRG BR008 (HERO, NCT05705401), PRESCISION (NCT02653755), PRIMETIME (ISRCTN41579286), and EXPERT (NCT02889874) are evaluating de-escalation of RT in favor of ET. This suggests a deep need for trials that mitigate RT toxicities.

Preoperative RT (PRT), with its smaller target volumes and accelerated regimens addresses many of the above concerns. Published data from Maryland^15^, Duke^16^, Preoperative Accelerated Partial Breast Irradiation (PAPBI)^17^, UCMC (Ablative)^18^, Sunnybrook (SIGNAL)^19^, and UTSW^20^ have demonstrated that PRT is well tolerated and offers excellent local control rates. However, the tolerable RT dose that results in cure is unknown. Our trial aims to answer this question, and here we report our interim results.

## Materials and Methods

### Study design

Our novel IRB approved (CR22030170-007), HIPAA Compliant study aims to explore the feasibility, safety, and tolerability of this Phase Ib dose-escalated PRT in T1-T2N0 luminal breast cancer (NCT05464667). The trial has two parts: a dose-escalation part evaluating PRT in four cohorts to determine the maximum tolerated dose (MTD), and a dose-expansion part where an additional 6 patients are enrolled at the MTD for additional safety and efficacy. In the first cohort, patients received a standard dose of 30 Gy in 5 fractions every other day to the Gross Tumor Volume (GTV) and Planning Target Volume (PTV). Each cohort dose-escalates the GTV by 5 Gy, except cohort three to four which escalates by 10 Gy. As such the GTV in cohorts 2 and 3 receive 35 Gy and 40 Gy, respectively, with interim analysis following completion of enrollment on cohort 3 for toxicity analysis. A total of four planned cohorts, with a maximum tested dose of 50 Gy to the GTV, are proposed in this study. BCS was performed within about one week (five to ten days) of completion of PRT.

### Eligibility

Post-menopausal women with unifocal T1-2, estrogen receptor positive, HER 2 negative invasive breast cancers were eligible for enrollment. Those with prior RT to the index breast were not allowed. Those with prior cancers, including contralateral breast cancers, were allowed, if treatment was completed 5 years prior to this diagnosis.

### Planning and Treatment

#### CT Simulation and Target Delineation

Patients underwent CT (GE LightSpeed RT; GE Medical System, Milwaukee, WI) scanning with 1.25 mm slice thickness either in the prone position on a CDR Procline prone breast board (CDR Systems Inc, Calgary, AB, Canada) or in the supine position with a usual breast board with 5 to 10 degrees of incline, which was placed on top of the CT table and treatment couch.

The breast MR images (if obtained before planning CT simulation) and planning CT images were imported into the Eclipse treatment planning system (TPS) (Varian Medical Systems, Inc., Palo Alto, CA). MR and CT image sets were registered to align the biopsy marker and soft tissue around the tumor area using manual rigid-body registration. All structures were contoured on the CT. GTV was identified as the area of enhancement on contrast-enhanced MRI with the assistance of a radiologist specializing in breast imaging. The GTV was dose-escalated, per protocol, depending on cohort. The CTV was expanded 10 mm from the GTV and conventionally named by its assigned dose of 30 Gy. The CTV was cropped 5 mm out of the skin and at the chest wall. The PTV was expanded 3 mm from the CTV and named by its assigned dose of 30 Gy. The PTV was cropped 5 mm out of the skin and at the chest wall. The normal tissue volumes contoured, per the RTOG breast contouring atlas, included the following: chest wall, skin uninvolved normal breast, contralateral breast, thyroid, ipsilateral lung, contralateral lung, and heart.

#### Treatment Planning and Delivery

All plans were done with either multi beam static IMRT or VMAT with partial arcs using the anisotropic analytical algorithm (AAA version 16.1) for dose calculation. Beam isocenter was set at the geometrical center of GTV. All plans had gantry and collimator set to avoid contralateral breast and heart based on beams eye view (BEV) in TPS. No beams entered from the back or side of a patient through a lung. All beams entered directly through the ipsilateral breast to reach to the target effectively. In general, the gantry angles were set to be spread as much as possible (15° to 30°) among beams within the limited range. Varian Halcyon (Varian Medical Systems) 6-FFF photon beam with 800 MU/min dose rate was used for treatment plan dose calculation. Dose volume constraints were followed per the protocol as followings (NCT 05464667). IGRT was employed prior to each fraction, with CBCT, to assist with localization and alignment of the target volumes using fiducial markers/biopsy clips.

### Breast Imaging/MRI

Breast imaging was obtained as per standard of care as part of the diagnostic workup for breast cancer and included bilateral mammography with tomosynthesis, targeted ultrasound of the breast malignancy and ipsilateral axilla, and pre-treatment MRI to assess disease extent. Breast MRI was also recommended following PRT to ascertain treatment response prior to definitive breast-conserving surgery (BCS). Breast MRI was performed on 1.5T Signa HDxt or Artist platform (GE Healthcare), or 3T Magnetron or Prisma platform (Siemens Healthineers) using a dedicated prone 8-or 16-channel breast coil. Bilateral axial short-tau inversion recovery images were obtained, followed by dynamic T1-weighted fat-suppressed images both prior to and three times following the intravenous administration of a weight-based dose of gadoteridol (0.1 mmol/kg) or gadopiclenol (0.05 mmol/kg) (Bracco Diagnostics, Inc.) gadolinium-based contrast. Delayed post-contrast T1-weighted fat-suppressed sagittal images were also obtained. Maximum intensity projection images were created from the first subtractions, and kinetic analysis was performed using DynaCad (Philips) software. All MRI examinations were reviewed by a breast imaging radiologist (WB) with 33 years of experience interpreting breast MRI.

### Pathology Evaluation and Magee Equations Calculation

Pre-therapy 14-g or 9-g core biopsies were processed and evaluated during routine pathology examination. All invasive carcinomas were evaluated for estrogen receptor (ER), progesterone receptor (PR), HER2, and Ki-67 via immunohistochemistry (IHC) and interpreted according to American Society of Clinical Oncology/College of American Pathologist (ASCO/CAP) guidelines. (PMID: 31928404; PMID: 29846122) Additionally, semiquantitative scoring was also performed for ER and PR using the histologic score (H-Score) method. H-score is calculated as the sum of intensity of staining times the proportion of cells staining and has a dynamic range of 0 (negative, no staining) to 300 (diffuse strong staining). HER2 IHC 2+ cases were further evaluated by fluorescence in-situ hybridization (FISH) and classified as positive or negative using the ASCO/CAP criteria. Ki-67 proliferation index was calculated as percent positive tumor cells in the entire tumor in the core biopsy sample. ER, PR, HER2, and Ki-67 results were then used to calculate the Magee Equation 3 (ME3) scores. Magee Equation 3 (ME3) is one of the 3 multivariable models that have been shown to have prognostic and predictive value in ER+/HER2-negative breast cancers. (PMID: 28548119; 32661297; 32203092) Magee Equation model was originally created to estimate the onctotype Dx recurrence score but has been shown to have chemopredictive value in the neoadjuvant setting as a stand-alone test. (PMID: 18360352; 23503643) In the neoadjuvant setting, a score of less than 18 has been shown to have a pathologic complete response (pCR) rate to chemotherapy of 0%, score of 18 to 25 with pCR rate of <5%, score of >25 but <31 with pCR rate of 10-15%, and score of 31 or higher with pCR rate of 35-40%. (PMID: 18360352; 23503643) The ME3 score based on pre-therapy receptor results was calculated on all cases.

The post-therapy resection specimen was processed according to standard pathologic examination for neoadjuvant therapy cases. Percentage tumor volume reduction in the breast was calculated based on comparison of the cellularity of the post-therapy tumor bed with the invasive carcinoma cellularity in the pre-therapy core biopsy (whenever available).

### Quality of Life (QOL) and Cosmetic Assessments

QOL and cosmesis were assessed using multiple measurements including: 1) Breast Cancer Treatment Outcome Scale (BCTOS self-administered questionnaire) 2) European Oranization for the Research and Treatment of Cancer Qualify of Life Questionnaire BR23 (EORTC QLQ BR23) 3) Breast Cosmesis Assessment – Harvard Scale 4) Digital Photography. This is to be performed at the following intervals: within 14 days prior to PRT; 1 month following PRT; 6 months following PRT; years one to five following PRT.

### Statistical Considerations

Continuous variables were summarized with their mean, standard deviation, median and range. Categorical variables were summarized with their proportion and percentage, along with the corresponding exact 95% confidence interval. For each continuous outcome, the pre- versus post-treatment difference was tested with the paired t-test. The comparison between groups was done with the two-sided two-sample t-test. The correlation between ordinal outcomes was tested with the Spearman rank correlation coefficient.

### Blood-and Radiomic Based Biomarkers

Blood was drawn for research purposes to assess cancer response to PRT through blood-based biomarkers. Assessment of cytokine stimulation via the cytokine reporter cell line was evaluated. This was performed within 14 days prior to PRT, within 7 days of completion of PRT, and 1 month following PRT.

Radiomic analysis was performed on available imaging for those who received pre- and post-PRT bilateral breast MRIs.

## Results

In this phase IB study, three patients were enrolled in cohorts 1, 2, and 3 for a total of 9 patients enrolled. Table 1 lists detailed demographics for those enrolled. Eight of the nine participants identified as white and non-Hispanic/LatinX, and one patient as East Asian, non-Hispanic/LatinX. None of the enrolled patients identified as either African/African American or Hispanic/LatinX. All patients were female and post-menopausal, with median age 60 years (range 51 to 71 years old). Enrolled patients had clinical T1-2,N0 disease and were noted on final pathology to have ypT1b to T1c disease, and all were invasive ductal carcinoma, estrogen and progesterone-receptor positive, HER2 immunohistochemistry-negative. One participant was noted to have isolated tumor cells in a sentinel node and all nine had otherwise negative sentinel node biopsy. Clear margins were achieved in all nine participants. No further RT was indicated as per standard of care (e.g. for positive margins, micro/macrometastatic node positive disease, etc) following BCS.

Oncotype testing was performed in four patients and scores ranged from 6 to 24 (median of 12.5). No patients received chemotherapy, and all patients were recommended ET following completion of BCS.

### Toxicity

While on treatment, acute toxicity grades ranged from zero to one for all nine participants (median of 0 as only 3 patients had grade 1 toxicity), with the most common toxicity being radiation dermatitis. At the time of one-month follow up from completion of PRT, and following completion of BCS, acute toxicity scores ranged from zero to two (median of 1, with only 1 person with grade 2 toxicity which was lymphedema which was in cohort 1). None of the nine patients developed ≥ grade 2 acute skin toxicity (which was the dose-limiting toxicity for this trial). No patients in cohort 3 had ≥ grade 2 acute toxicity overall at one month following completion of PRT.

### Quality of Life and Cosmetic Outcome

At one month following PRT, 7 participants opted to complete the BCTOS and EORTC surveys. At 6 months, 5 and 6 participants opted to complete the BCTOS and EORTC surveys, respectively. All 9 participants had cosmetic analysis at 1-month post-PRT using the Harvard scale. At one month and six months after completion of PRT, no differences were noted in either quality of life or cosmetic outcome as noted on the BCTOS, EORTC QLQ BR23, and Harvard Scales (p≥0.05). Digital photography evaluation will be performed at time of final analysis.

### Pathologic Correlation

No patients were noted to have a pathologic increase in size of their primary malignancy compared to initial imaging. Post op path showed tumor sizes ranging from 0.1 cm to 1.9 cm (median 1.0 cm).

For most patients, the final pathology size was reflective of the maximum tumor size seen on MRI. Those enrolled in cohorts 1 and 2 had little change in their final tumor size compared to that noted on MRI. Interestingly, two of the three patients in cohort 3 experienced significant shrinkage in their final pathology compared to MRI measurement. For one patient (C3-01), her tumor measured 1.3 and 1.5 cm in pre- and postRT MRI and 0.55 cm on final pathology. For the second patient (C3-02), her tumor measured 0.9 and 0.7 cm on pre- and postRT MRI and 0.1 cm on final pathology. Of note, the third patient (C3-03) had a 0.8 cm tumor on pre- and post-RT MRI and a 1.0 cm tumor on final pathology.

No patients enrolled in cohorts one to three achieved a pathologic complete response (pCR). One participant in cohort four achieved a pCR, and this will be included in the final analysis.

### Volumetric Changes of Index Cancer on MRI

Only 6 of 9 participants had both pre- and post-PRT MRIs. Of the 7 with pre-PRT MRIs, all were noted to have an irregular mass on preRT MRI. Six patients had a pre and post RT MRI showing an irregular mass with homogenous or heterogeneous internal characteristics on pre-and post MRI. Kinetics seen generally ranged from persistent, plateau, washout, or mixed pretreatment. The maximum dimension ranged from 0.6 to 2.1 cm on preRT MRI. For those who had a postRT MRI, the mass sizes ranged from 0.7 to 2.0 cm.

One patient in cohort three had an increase in size of the primary malignancy on MRI, while three patients had a decrease (one in cohort two and two in cohort three), and two patients (one in cohort one and one in cohort two) had no change on MRI. For one participant in cohort 3, new increased non-mass enhancement was appreciated on post-PRT MRI that was not seen on pre-PRT MRI. This corresponded to extensive DCIS noted on surgical resection specimen. Figure 1 shows representative MRI images from patients in cohorts one to three pre- and post-PRT.

### Blood- and Radiomic-based biomarkers

All 9 participants has blood-based biomarker analysis. Cytokine expression was measured about 2 weeks prior to PRT as well as one week and one month after PRT. A significant increase in the activation of the canonical NF-κB pathway via increased TNF-α and IL-1β was appreciated with dose-escalated PRT to 40 Gy (cohort three) (p≤ 0.001).

Radiomic analysis of pre- and post-PRT MRIs was performed. Of significance was a decrease in elongation values (p=0.0409) and increase in original first order 10 percentile values (p=0.0390).

This suggests that the tumor size reduce (was less elongated) and had more necrosis/inflammatory changes, respectively.

Tissue biomarker analysis will be conducted once all four cohorts have completed enrollment.

## Conclusions

Our interim analysis shows that dose-escalated PRT to the GTV is safe, feasible, and well-tolerated compared to historical controls. While on treatment, patients did not experience ≥grade 2 toxicity, and one-month following completion of PRT and BCS, no patients had ≥grade 3 toxicity. No wound complications were reported.

A large benefit of PRT, when compared to post-operative RT is the smaller treatment volumes. This will potentially result in lower rates of acute and long-term toxicities, especially when PRT is only dose-escalated to the GTV. Additionally, there is less integral dose to the remainder of the breast as well as normal tissues of heart, lung, chest wall with PRT than standard partial breast or whole breast RT. In light of this, PRT could enable more people to undergo partial breast irradiation as it would be easier to achieve planning constraints. Furthermore, BCS removes the irradiated area, thereby potentially reducing long-term fibrosis. Finally, PRT does not preclude the ability for oncoplastic surgical approaches as RT is completed prior to BCS.

### Imaging and Pathologic Response

While imaging response was variable, following PRT, no patients were noted to have achieved a pCR. However, with dose-escalation, two of three patients enrolled in cohort three did have an impressive partial response. Data from Duke show a pCR rate of 11% when surgery is completed in 10 days^16^. Increased response rates are noted with increased time from PRT to surgery. At Maryland, pCR was 15% if surgery followed in 21 days, while at UCMC, pCR rates were 33% and 48% when surgery followed at six and eight months, respectively. In dose-escalated single fraction PRT to the PTV, (with BED of 40 Gy in 5 fractions at the highest dose level), no patients at UTSW experienced a pCR. However, pathologic response to dose-escalation to just the GTV, beyond 40 Gy in 5 fractions has not been reported, but was noted in the first participant enrolled in cohort four (50 Gy). This participant was noted to have a clinical complete response on post-PRT MRI and pCR on final pathology following BCS. Continued enrollment and evaluation of those in cohort four is warranted. By achieving a pCR, the role of BCS, and perhaps even intensified risk-reducing systemic therapy is called into question with further exploration and study required.

### Quality of Life and Cosmetic Outcome

At one and six months following PRT, no differences in QOL or cosmetic outcome were appreciated. These findings are promising as they suggest that those who receive radiation pre-operatively do not do worse than those who receive it standardly in the post-operative setting. Continued follow up for QOL and cosmesis is warranted to determine the long-term effects of PRT.

### Biomarker Response to PRT

To our knowledge, the investigators at Duke reported tissue-based but not blood- or radiomic-based biomarker analysis, suggesting that this is a critical area of inquiry to understand the systemic response of PRT^21^. Our study is notable for significant cytokine stimulation resulting in activation of the canonical NF-κB pathway following delivery of PRT. This suggests important systemic immune response from intact breast cancer to high-dose radiation. Comparison of the tissue microenvironment and biomarkers seen on core biopsy and post-PRT BCS specimen will assist with our understanding of such a notable effect.

Furthermore, while limited tumoral changes were appreciated radiologically on MRI in cohorts one to three, significant changes were seen in radiomic biomarkers notable for change in cancer shape and cellular density. Additional evaluation for those in cohort four (with one participant noted to have a clinical complete response) will be critical. Comparison to final pathology will be important to allow prediction of cancer response to PRT.

### Clinical Trial Demographics

Demographically, participants are primarily of white race. Efforts to increase access to those across racial demographics and rural/urban location are underway. Support has been secured from the Pittsburgh Foundation to assist with increasing community engagement. Furthermore, this trial sits within the NRG Health Access Committee Rural Health Special Interest Group as a mechanism to advance trial enrollment for all populations. There is hope that by improving engagement, this study would reduce barriers to care by offering just five fractions of PRT, thereby improving oncologic outcomes. ^22,23^

### Future Directions

Dose-escalation of PRT to 40 Gy to the GTV appears safe, feasible, and well-tolerated. In light of this, the trialists will move forward with subsequent cohorts of this study. Further reports of tissue and radiomic biomarkers, as well as longer-term toxicity, will be presented in future analyses. ^24^

## Supporting information

Demographics

MRI

## Data Availability

All data produced in the present study are available upon reasonable request to the authors

