## Supplementary material for "Interim Analysis of Dose-Escalated Preoperative Breast Irradiation: A Phase Ib study": Demographics

**Table 1. Demographics**

| Characteristics | n(%) |
|---|---|
| **No. of patients** | 9 |
| **Age**: Median (range) | 60 (50-71) |
| **Race** | |
| East Asian | 1 (11%) |
| White | 8 (89%) |
| **Ethnicity=Hispanic/Latino (Y/N)** | |
| N | 9 (100%) |
| **Clinical stage** | |
| T1bN0 | 6 (67%) |
| T1cN0 | 2 (22%) |
| T2N0 | 1 (11%) |
| **Pathological stage** | |
| T1bN0 | 5 (56%) |
| T1cN0 | 2 (22%) |
| T1cN0(i+) | 1 (11%) |
| pT1miNx | 1 (11%) |
| **IDC or ILC pre-RT (1=IDC, 2=ILC, 3=Both)** | |
| 1 | 8 (89%) |
| 3 | 1 (11%) |
| **IDC or ILC post-RT (1=IDC, 2=ILC, 3=Both)** | |
| 1 | 9 (100%) |
| **Nottingham grade pre-RT** | |
| 1 | 4 (44%) |
| 2 | 3 (33%) |

| **Characteristics** | **n(%)** |
|---|---|
| 3 | 1 (11%) |
| Not Performed | 1 (11%) |
| **Nottingham grade post-RT** | |
| 1 | 3 (33%) |
| 2 | 6 (67%) |
| **ER H score pre-RT**: Median (range) | 300 (235-300) |
| **PR H score pre-RT**: Median (range) | 268 (3-300) |
| **HER2 Immunohistochemistry pre-RT** | |
| 0 | 2 (22%) |
| 1 | 5 (56%) |
| 2 | 2 (22%) |
| **HER2 amplified (1=yes (0, 0%), 2=no, 3=not performed) pre-RT** | |
| 2 | 2 (22%) |
| 3 | 7 (78%) |
| **Ki67 percent pre-RT**: Median (range) | 7.5 (2-25) |
| **Oncotype score pre-RT (n=4)**: Median (range) | 12.5 (6-24) |
| **Magee score pre-RT**: Median (range) | 13.3 (9.5-21.4) |
| **Max dimension size (cm) pre-RT (n=7)**: Median (range) | 1.1 (0.6-2.1) |
| **Max dimension size (cm) post-RT (n=7)**: Median (range) | 1.4 (0.7-2) |
| **Max tumor size on final path (cm)**: Median (range) | 1.0 (0.1-1.9) |
