## Supplementary material for "Interim Analysis of Dose-Escalated Preoperative Breast Irradiation: A Phase Ib study": MRI

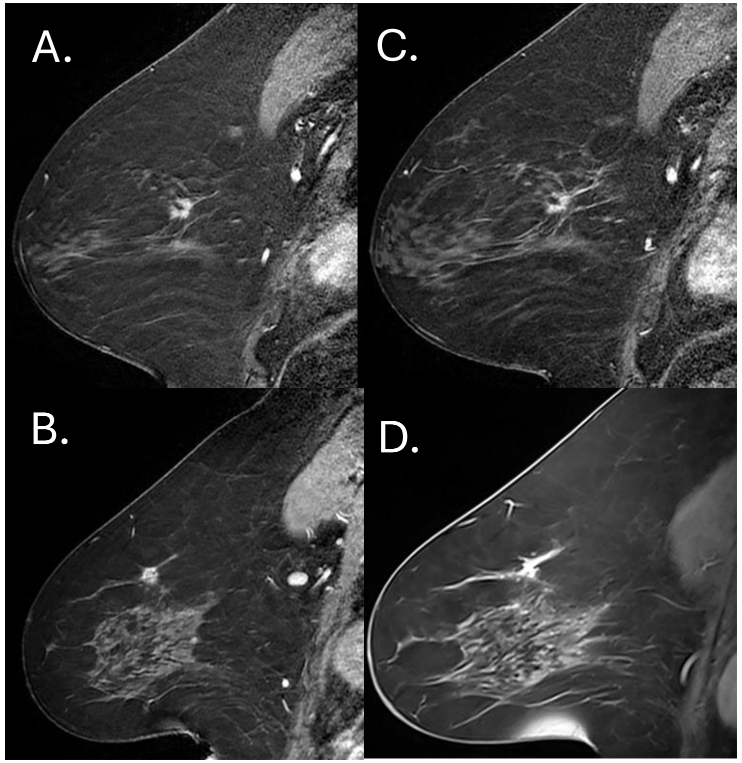


Figure 1. Sagittal views of MRI showing the intact breast cancer prior to preoperative radiation (PRT) for cohorts two (35 Gy, A) and three (40 Gy, B). Sagittal views of MRI showing limited imaging cancer response after PRT for the same participant in cohorts two (C), and three (D).
